# Regional variation and urban-rural differences in partner similarity: a nationwide cohort study

**DOI:** 10.64898/2026.06.23.26356303

**Authors:** Eetu Soini, Kateryna Golovina, Kimmo Suokas, Mai Gutvilig, Marko Elovainio, Markus Jokela, Christian Hakulinen

**Affiliations:** Department of Psychology, Faculty of Medicine, University of Helsinki; Population Research Institute, Väestöliitto ‒ Family Federation of Finland, Helsinki, Finland; Finnish Institute for Health and Welfare, Helsinki, Finland

## Abstract

Although romantic partners tend to resemble each other on many characteristics, the geographical processes underlying partner similarities remain poorly understood. Using Finnish nationwide registry data from cohabiting or married partners (N = 1,500,204 couples; partnerships were formed between 1990-2023), we examined regional differences in partner similarity in mental disorders, educational attainment, and adolescent school performance. We also analysed geographical variation in partner similarity within three major cities. Accounting for local demographic composition of potential partners attenuated the partner similarity from r=0.43 to r=0.34 for highest obtained educational attainment, but increased partner similarity in any mental disorders from r=.40 to r=.42. In urban municipalities partners were more similar in educational attainment, but less in mental disorders, compared to more rural regions. We found no clear within-city variation in partner similarity. These findings highlight the role regional demographic composition plays in partnership formation and suggest different partnering dynamics depending on societal organization.

## Introduction

Partner similarity is a widely observed phenomenon. Partners tend to be more alike in ethnicity, political values, religion and educational attainment ^1–3^, intelligence and personality ^4,5^, as well as physical and mental health ^6–9^. Recent findings have highlighted more complex associations between traits. Mental disorders seems to be affected by indirect assortment, meaning that assortment may happen on traits that are correlated with mental disorders. Assortment may occur on psychiatric vulnerability ^7,9,10^, so that partners may resemble each other in the latent vulnerability, but may not share the same diagnosis. Or assortment could happen on traits such as education, so that similarity in mental disorders emerges from assortment on education ^9^. These more complex patterns make it more difficult to identify the traits driving the assortment. Furthermore, recent findings on partner similarity in educational attainment broadly agree that similarities cannot be explained solely by direct assortment. Kinship studies systematically report higher-than-expected similarities in educational attainment between genetically and non-genetically (in-laws) related relatives ^11,12^, highlighting that environmental factors role in assortment. Complementary findings come from sibling and twin studies focusing on partners of relatives, where both phenotypic assortment, but also shared environmental factors seem to contribute to partner similarity in educational attainment ^13,14^.

While assortment can operate through individual preferences for partners, it can also operate through mechanisms that facilitate interaction between people. One such mechanism is stratification, i.e., the division of populations based on certain traits. This may be due to the geographic clustering of traits or social stratification, that is, people more likely to interact with members of their social groups. Geographic proximity has influenced partnering throughout human evolution, contributing to genetic differences both between and within continents and countries ^15,16^. This reflects the tendency for individuals to form partnerships with people from nearby areas, a process often referred to as isolation by distance ^17^. While differences in genetic structure largely reflect historical migration flows and geographic barriers, geographic proximity continues to influence partnering patterns in modern societies. Studies of propinquity have shown that spouses tend to originate from nearby locations. For example, early studies in the United States found that many married couples had lived within a few city blocks of each other prior to marriage ^18^ with similar findings reported later in the U.S. (Clarke, 1952), in the Netherlands ^20,21^, and in Finland, where, historically, a large share of marriages occurred between partners from the same municipality (Haavio-Mannila, 1964). While urbanization and modernization are hypothesized to affect partner selection (Maas & Zijdeman, 2010), geographic proximity remains important, as physical closeness increases opportunities for interaction, and interactions facilitate partnership formation (Kalmijn, 1998; Kalmijn & Flap, 2001).

From the social stratification perspective, barriers do not need to be physical. Within urban centers where physical distances become smaller, stratification is more likely based on social characteristics. Blau and Schwartz ^22^ have proposed that regions differ in their dynamics that lead to partner similarity due to differences in population size, demographic composition, but also how social circles are organized. First, within larger populations the pool of potential mates is larger even for small subgroups, increasing the chances of finding a partner with a matching level of a trait. Second, urban centers are characterised by greater heterogeneity, which can facilitate interactions beyond one’s own social class, religion, or ethnic background. Third, increasing population size and population density may lead to social interactions becoming more revolved around institutions such as schools and work places that contribute to greater stratification ^23–26^. These hypothesized processes imply that regional context may alter partner selection in distinct ways, leading to variation in the degree and mechanisms of partner similarity between urban and rural areas. These processes suggest competing predictions for how partner similarity should vary across the urban-rural gradient: greater institutional stratification in cities may increase educational assortment, while greater social heterogeneity may reduce assortment on other traits. Prior work has found that urban areas are associated with higher education assortment ^27,28^.

In this paper our aims are four-fold. First, we investigated how much partner similarity in educational attainment and mental disorders is affected by the geographical clustering of these traits. Second, we explored if indirect assortment on educational attainment would explain partner similarities in mental disorders when geographical clustering is accounted for. Third, we studied the urban-rural gradient in partner similarity in Finland, to see if partnering dynamics differ between regions. We expect that higher urbanicity would be associated with greater educational assortment, which reflects stronger institutional stratification, and potentially with lower assortment on mental disorders, reflecting greater social heterogeneity in urban regions. We further tested if this gradient is affected when partner similarities in mental disorders are adjusted for indirect assortment on education. Fourth, we tested whether partner similarity varied within three major Finnish cities.

## Results

### Study sample

The study population included 1 082 674 men and 1 085 150 women forming 1 500 204 partnerships (i.e., marriage or cohabitation) between the beginning of 1990 and end of 2023. Finland as a context is discussed in supplementary materials. Prevalences for any mental disorder and individual ICD-10 categories, and means and standard deviations for educational achievement, highest obtained education attainment, and 9^th^ grade GPA are presented in supplementary Table S1 for the sample as a whole, and by urbanicity category. Prevalence for any mental disorder at individual level was 14.3% and the highest prevalences from the specific mental disorders were observed for anxiety disorders (F40-F48, 6.8%), mood disorders (F30-F39, 5.7%), and substance use disorders (F10-F19, 3.3%). No major differences were observed between urbanicity categories though municipalities with the highest urbanicity rating had slightly lower prevalences, for example, any mental disorder most urban municipalities had prevalence of 13.9%, while least urban municipalities had prevalence of 15.9%.

Figure 1 shows the used urbanicity classification, the location of three major cities (Helsinki, Tampere, and Turku), and information on municipalities by urbanicity level. In Finland, the population has migrated towards southern cities, with three major cities (Helsinki = 696 000 Tampere = 259 000, Turku = 210 000) accounting for over a fifth of the whole population. In our sample, 59% of the couples’ first cohabitation occurred in the most urban municipalities. In the most urban municipalities, the median distance between partners last place of residence prior to cohabitation was 12km, which increased with decreasing urbanicity to 56km in the least urban municipalities, highlighting the role of geography in partner selection. Cumulative proportions for each urbanicity classification are presented in supplementary Figure S1.

**Figure 1.**
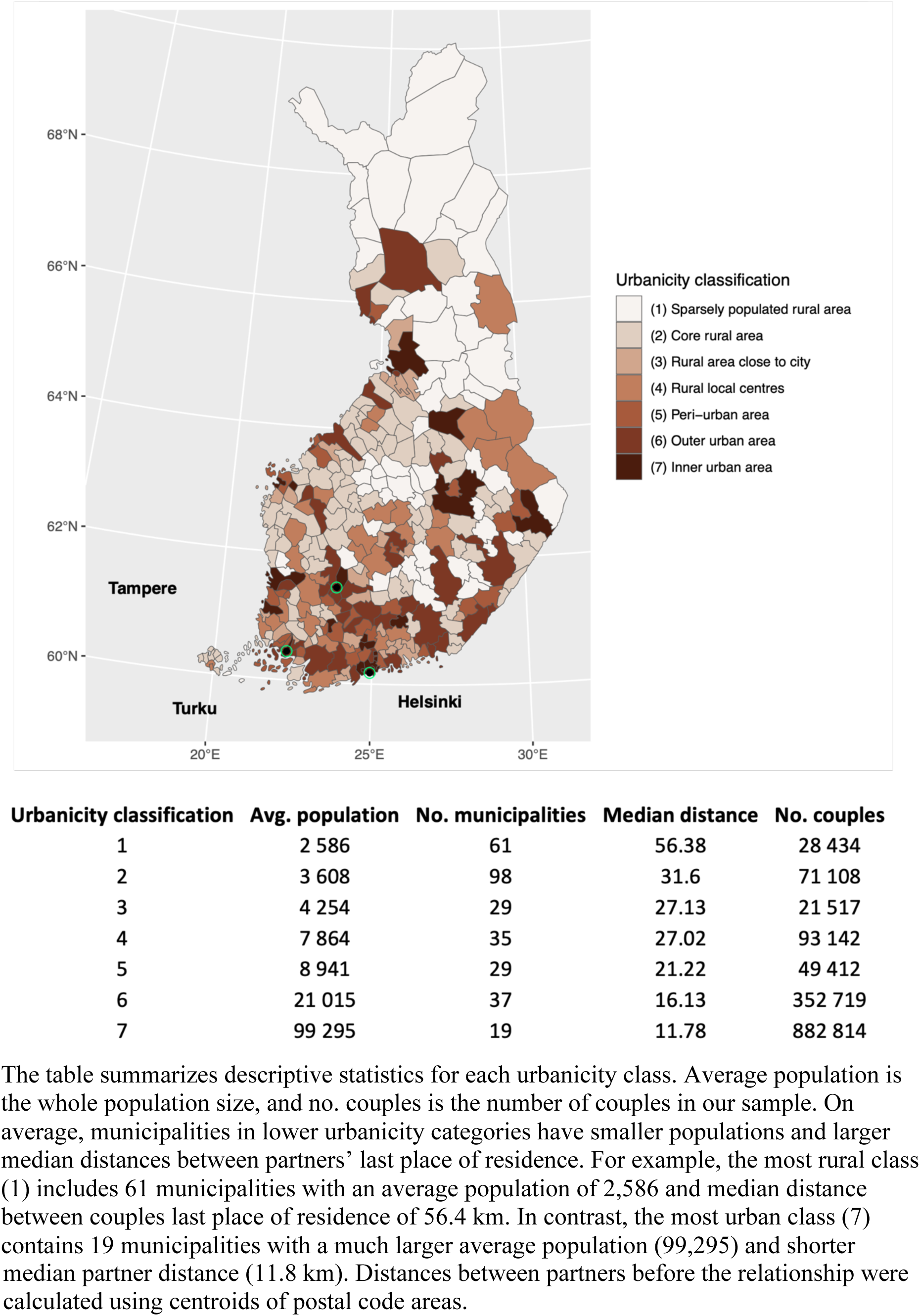
Map of urbanicity classification, descriptive statistics of municipalities and the location of three major cities (Helsinki, Tampere, and Turku)

### Partner similarity after accounting for local pool of potential partners

Figure 2 shows estimated partner similarity in mental disorders, educational achievement, highest obtained degree, and 9^th^-grade grade point average (GPA) calculated from the whole sample and aggregated estimate calculated within municipalities. A lower municipality-specific vs general-population correlation would imply that part of the assortment observed at general-population level can be attributed to spatial clustering. Thus, even if partnering is random, partners would be more similar because they are more likely to encounter others with similar traits within the regions they reside in. A higher estimate would imply that assortment within municipalities would be even higher when demographic composition of the municipality is accounted for. Highest similarity in the whole sample estimates was observed in intellectual disabilities (F70-F79), followed by schizophrenia spectrum disorders (F20-F29) and substance use disorders (F10-F19). When accounting for the composition of the local pool of potential partners (Fig. 2, estimates in light red), the estimates were on average 2.4% higher across diagnostic classes, with no major differences between diagnostic classes in the observed attenuation.

**Figure 2.**
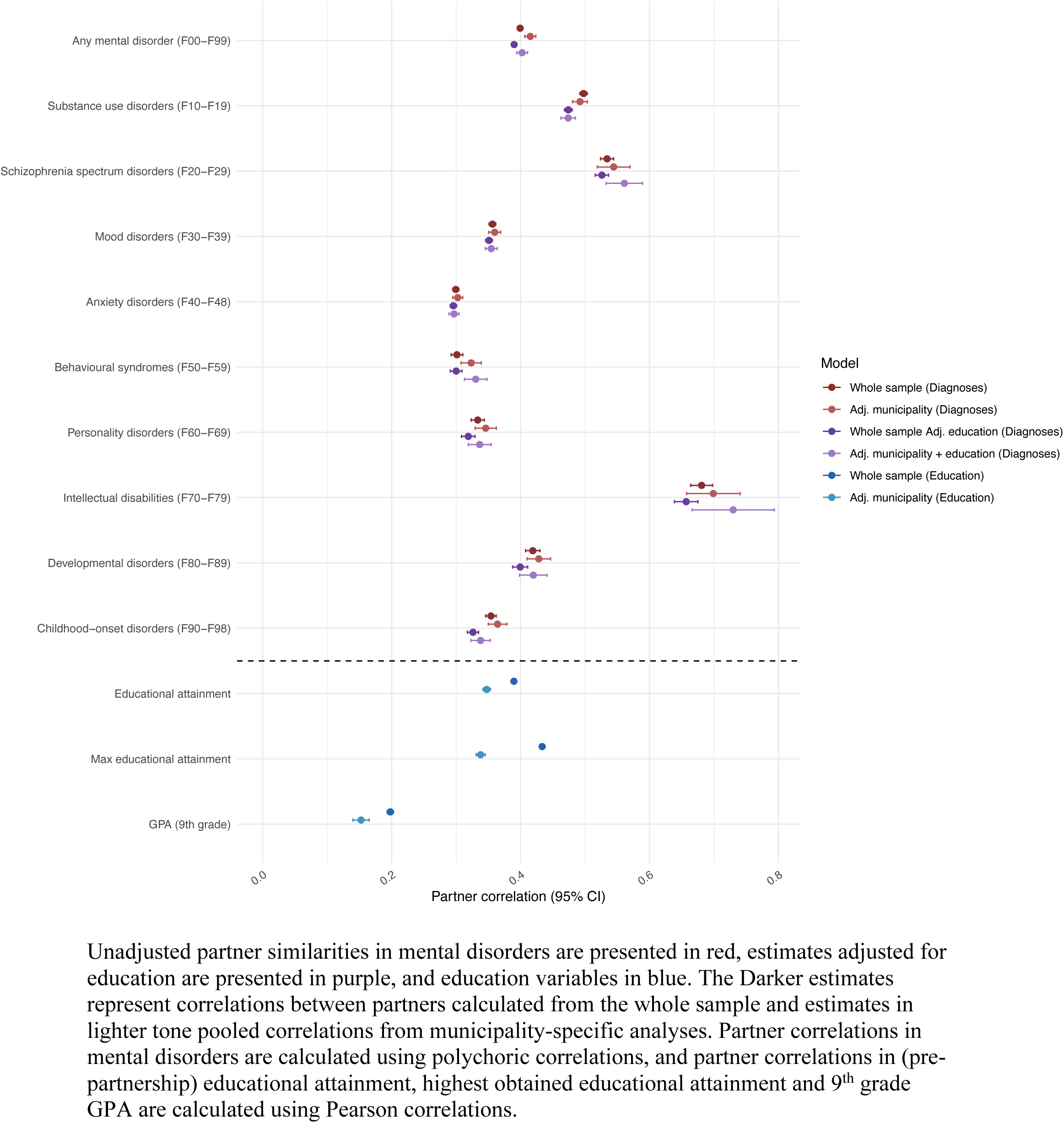
Comparisons between partner correlations in mental disorders and education from whole sample estimates and aggregated municipality estimates.

For pre-partnership educational attainment, the aggregated municipality-specific estimate was lower compared to the estimate calculated from the whole sample, indicating that spatial clustering accounts for 11% (whole sample: r=0.39 vs. municipality-specific: r=0.35). Highest obtained educational attainment showed a more pronounced attenuation of 22% (whole sample: r=0.43, municipality-specific r=0.34). Partner similarity in 9^th^ grade GPA was r=0.20, and attenuated to r=0.15 (24%) when accounting for the local pool of potential partners. Findings on partner similarity in education suggest that they may be especially affected by local demographic composition, potentially because educational institutes may act as one of the stratifying factors affecting partner similarity.

### Indirect assortment within regions

To assess whether earlier findings of indirect assortment between education and mental disorders ^9^ are partly attributable to geographic stratification, we adjusted partner similarities in mental disorders for pre-partnership educational attainment. If both educational attainment and mental disorders cluster geographically, observed partner resemblance may reflect shared regional distributions of these traits in addition to preference-based assortment. Estimates calculated from the whole sample were slightly lower for all specific mental disorder subchapters (avg. 3%) when adjusting for pre-partnership educational attainment (Fig. 2, estimates in dark purple), compared to the estimates unadjusted for education(Fig. 2, estimates in dark red). Comparing aggregated municipality-specific estimates adjustment for education made little difference in estimates (avg. attenuation 1%) (Fig. 2, light red vs. light purple).

### Urban-Rural differences in partner similarity

Results from the meta-regressions with urbanicity as a fixed-effect moderator are shown in Figure 3, and specific estimates are shown in Table 1. Most of the diagnostic categories did not show any trends with higher urbanicity (light red), with the exception of any mental disorder (F00-F99) (β = −.007), schizophrenia spectrum disorders (F20-F29) (β = −.029) and behavioral syndromes associated with physiological disturbances and physical factors (F50-F59) (β = −.025), all of which showed lower partner similarity in more urban municipalities (Table 1). Results were similar when using municipality-specific estimates adjusted for education (Fig 3., estimates in light purple), with differences by urbanicity more pronounced. Due to smaller samples in the most rural municipalities, meta-regressions did not converge for Intellectual disabilities (F70-F79) and developmental disorders (F80-F89).

**Figure 3.**
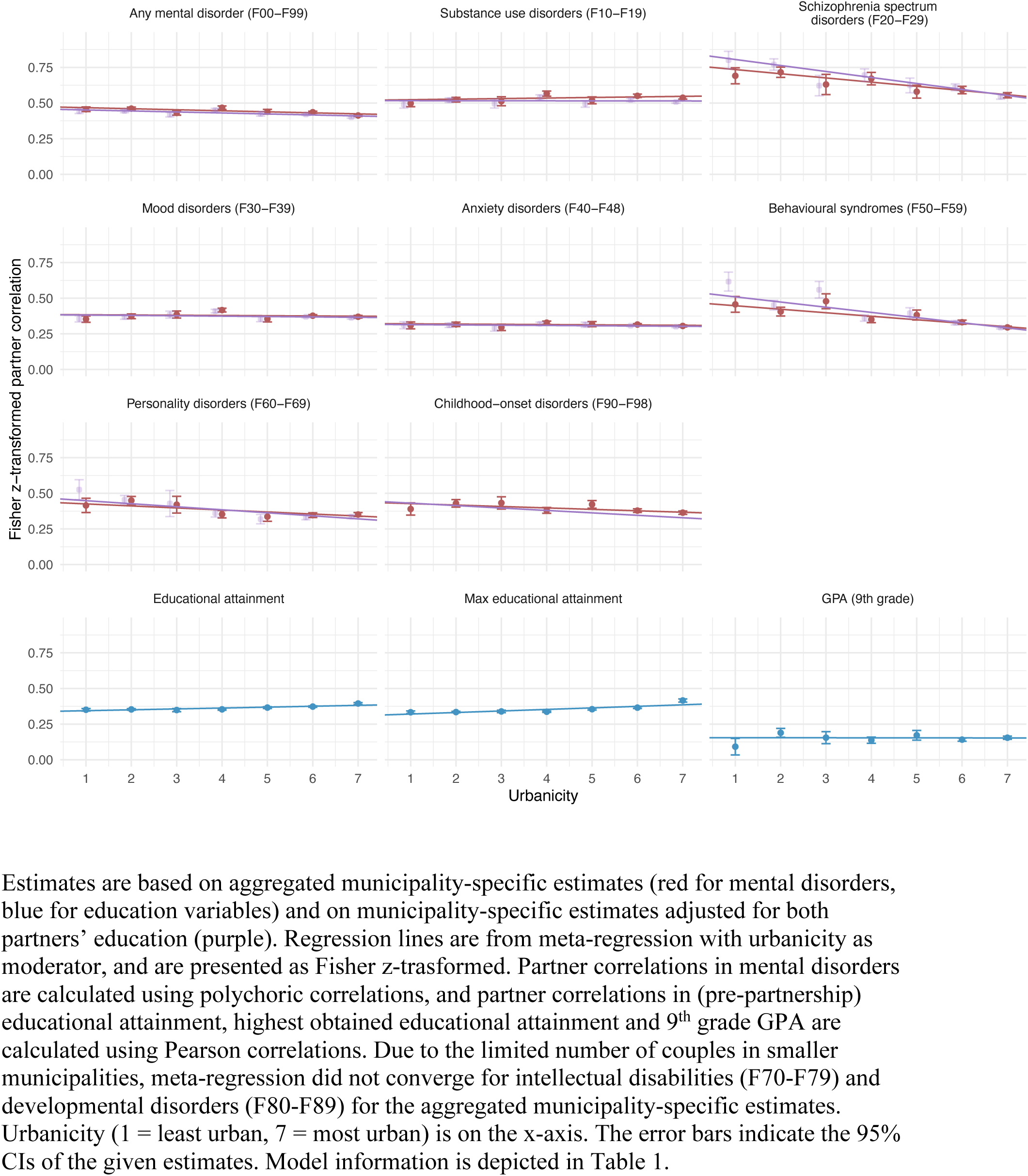
Meta-regression results with urbanicity as fixed-effect moderator

**Table 1.**
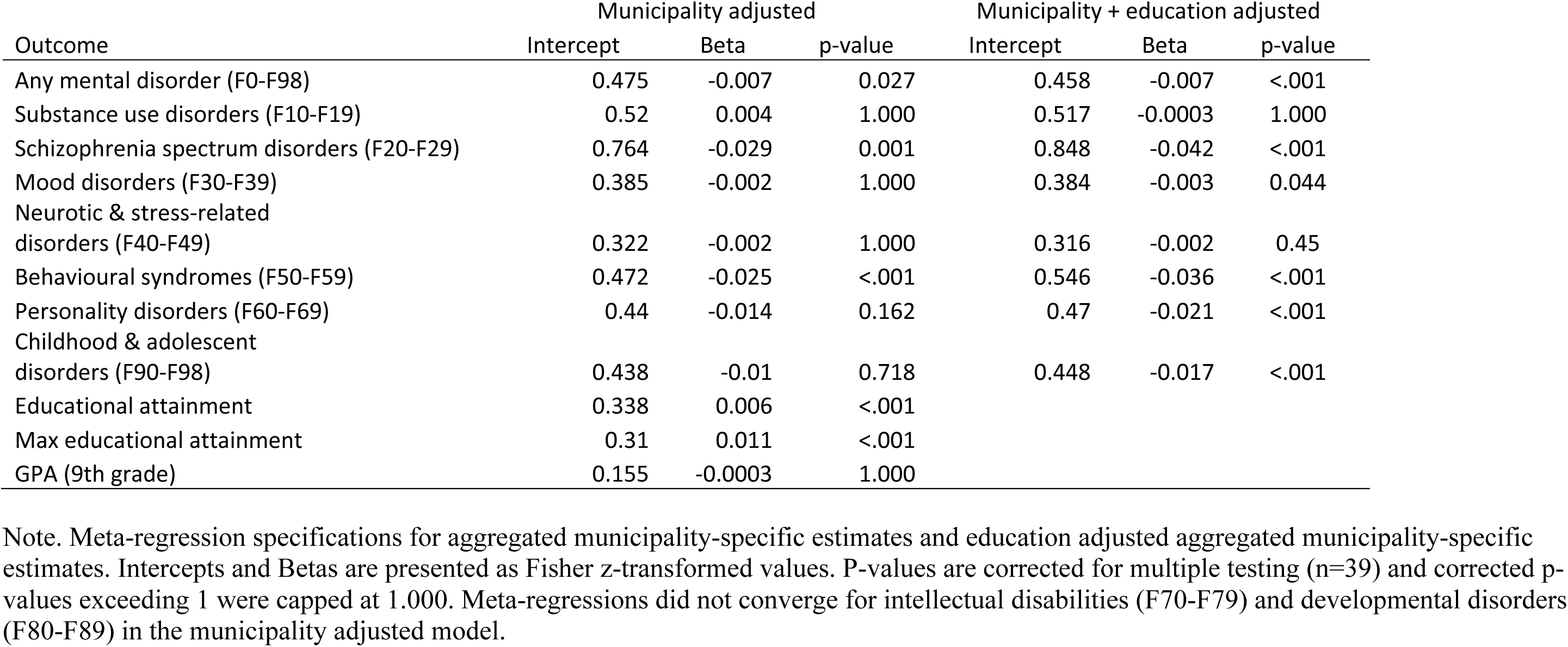
Estimates from meta-regressions with urbanicity as a fixed-effect moderator.

Higher partner similarity in pre-partnership educational attainment was observed with higher urbanicity (β = .006 per 1 level increase in urbanicity), and a stronger association for highest obtained educational attainment (β =.011). No differences were observed for 9^th^ grade GPA.

### Geographical proximity does not affect partner similarity at finer spatial scale

Lastly, we examined whether partner similarity differed across geographic division into subregions within three large municipalities. A map of Helsinki and its subdistricts is presented in Figure 4, and the accompanying table depicts the characteristics of each postal code area including total population, population density, median distance before partnership began, and analytical sample size. Maps of the two other examined cities, Turku and Tampere, are presented in supplementary figures S2 and S3. Population density varied substantially across areas, with the most densely populated central districts exceeding 8,000 inhabitants per km², while peripheral areas were less densely populated. Comparisons between municipality estimates (all couples formed within the municipality), and aggregated subregional estimates are depicted in Figure 5, and specific information about the models is presented in Table 2. Estimates calculated from municipalitys population and estimates aggregated from subregional estimates were similar in magnitude, and no meaningful differences were observed for either educational attainment or mental disorders. Likewise, population density did not show any meaningful trend with partner similarity in mental disorders or educational attainment (Table 2).

**Figure 4.**
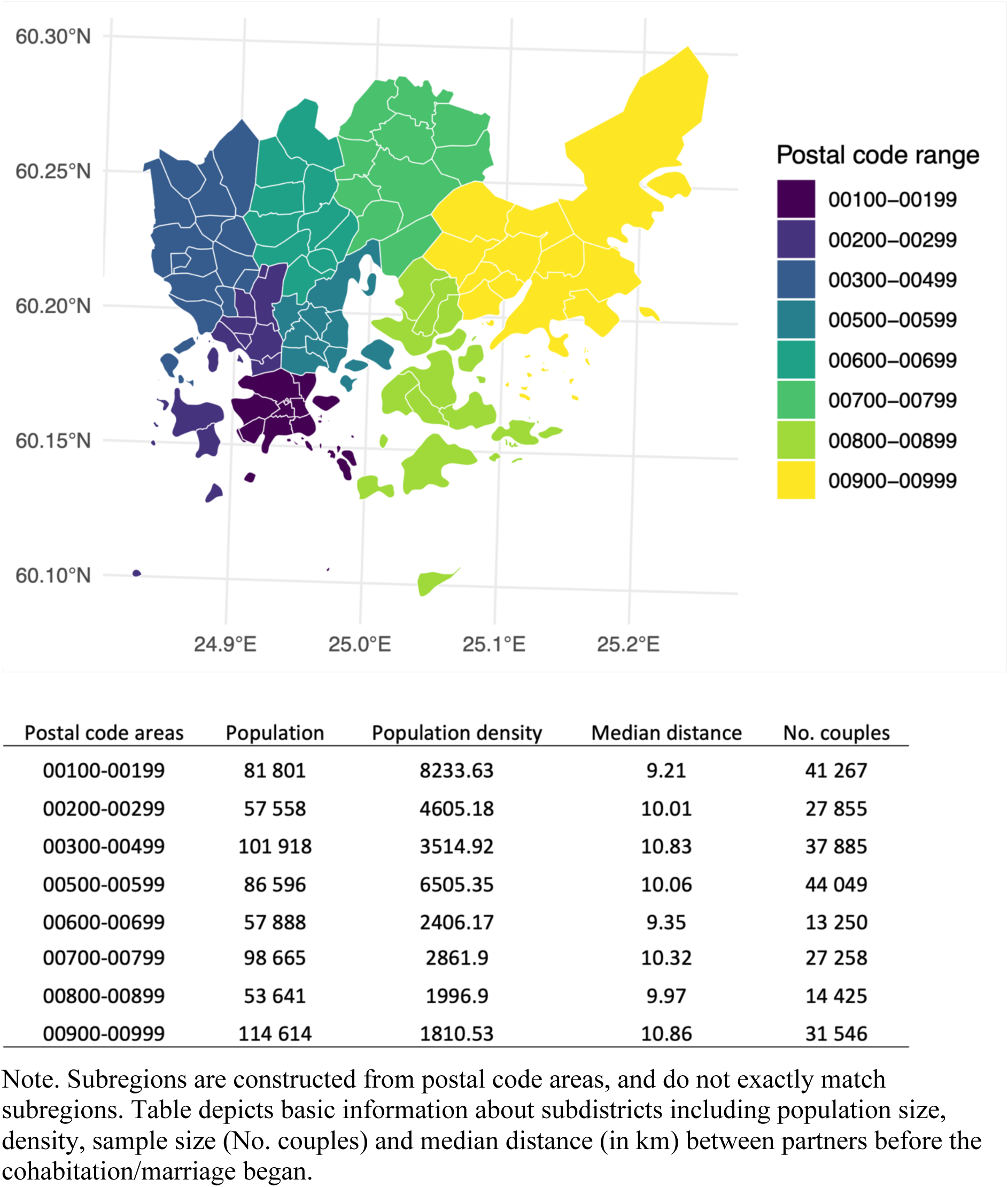
Map of Helsinki and descriptive statistics.

**Figure 5.**
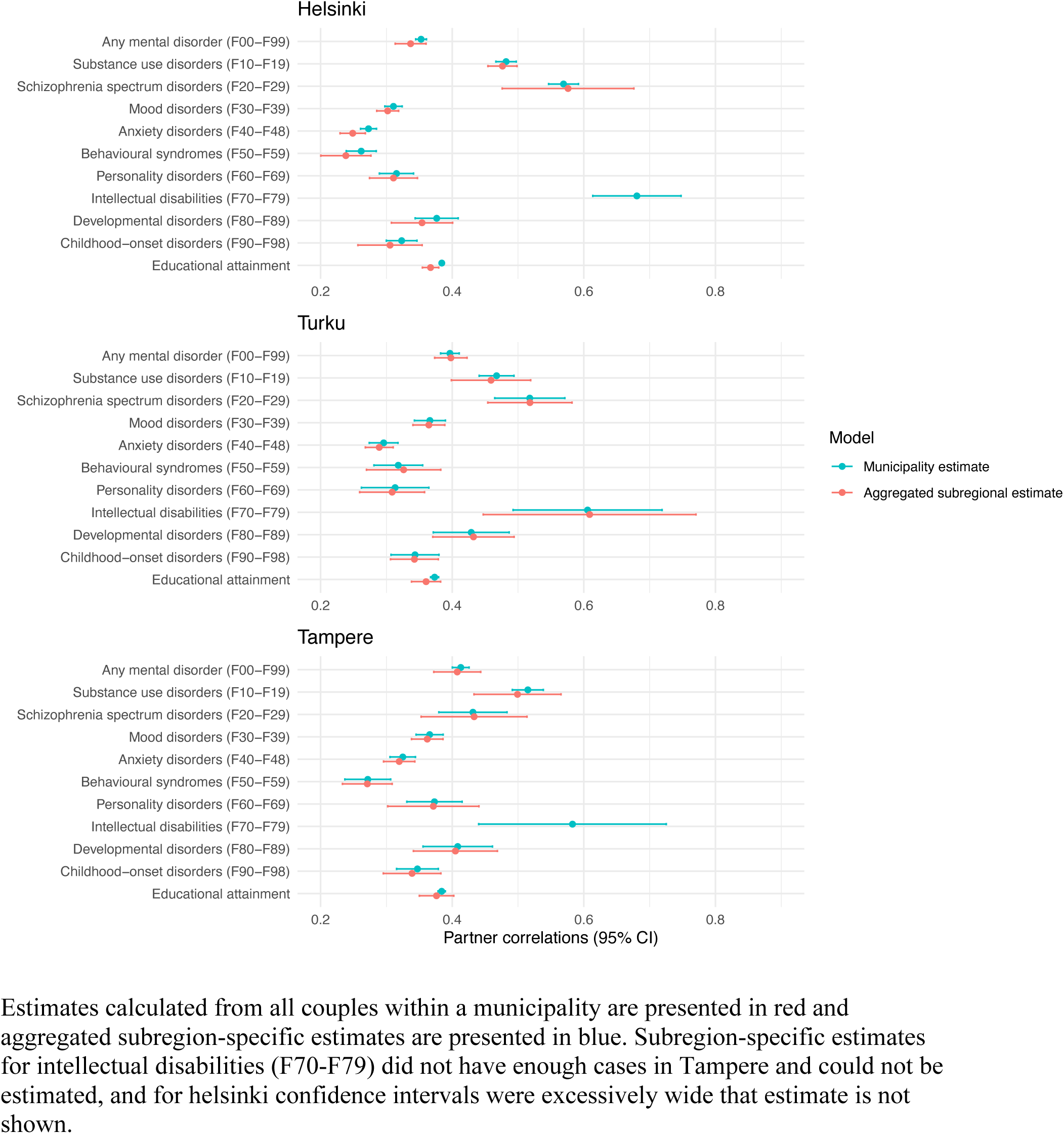
Comparisons of municipality estimates, and aggregated subregional estimates.

**Table 2.** Meta-regression estimates from subregional analyses with population density as the fixed-effect moderator.

|  | Helsinki |  | Turku |  | Tampere |  |
| --- | --- | --- | --- | --- | --- | --- |
|  | **B** | **P-value** | **B** | **P-value** | **B** | **P-value** |
| Any mental disorder (F00-F99) | -0.011 | 0.054 | -0.012 | 0.583 | -0.011 | 1.000 |
| Substance use disorders (F10-F19) | -0.007 | 1.000 | -0.029 | 1.000 | -0.010 | 1.000 |
| Schizophrenia spectrum disorders (F20-F29) | 0.002 | 1.000 | -0.012 | 1.000 | -0.066 | 1.000 |
| Mood disorders (F30-F39) | 0.0004 | 1.000 | -0.002 | 1.000 | -0.005 | 1.000 |
| Anxiety disorders (F40-F48) | -0.006 | 1.000 | 0.008 | 1.000 | 0.007 | 1.000 |
| Behavioural syndromes (F50-F59) | -0.012 | 1.000 | -0.016 | 1.000 | -0.02 | 1.000 |
| Personality disorders (F60-F69) | 0.008 | 1.000 | -0.001 | 1.000 | 0.033 | 1.000 |
| Intellectual disorders (F70-F79) | 0.019 | 1.000 | 0.003 | 1.000 |  |  |
| Developmental disorders (F80-F89) | 0.001 | 1.000 | -0.019 | 1.000 | 0.007 | 1.000 |
| Childhood-onset disorders (F90-F98) | -0.016 | 1.000 | -0.011 | 1.000 | -0.019 | 1.000 |
| Educational attainment | 0.0002 | 1.000 | 0.01 | 1.000 | 0.012 | 1.000 |

Betas and intercepts from subregions analyses for each of the three cities, presented as Fisher z-transformed. P-values adjusted for multiple testing ( $n = 39$ ) and corrected p-values exceeding 1 were capped at 1.000. Intellectual disorders (F79-F79) could not be estimated in Tampere due to a low number of cases.

### Sensitivity analyses

To test the robustness of our findings we examined whether the associations remained similar if we restricted the sample to those who had resided within the municipality for at least two years before the partnership began. Supplementary Figure S4 shows the results, and the prevalences for the subsample are presented in supplementary Table S2. Prevalences in the subsample were higher in more urban municipalities, and lower in the more rural municipalities (e.g., prevalence of any mental disorder in the most urban regions of the main sample = 13.9% vs. subsample 15.8%, and in least urban regions of the main sample = 15.9% vs. subsample 15.4%) leading to higher prevalences on the whole (prevalence on any mental disorder 14.4% vs. 15.8%). Thus, this subsample may capture certain characteristics that are negatively associated to residential migration, or associated with slower partnering. Estimates calculated from the subsample were, in general, of similar magnitude compared to the main analysis. Partner similarity for schizophrenia spectrum disorders (F20-F29) and intellectual disabilities (F70-F79) was markedly higher in the subsample than in the main analysis. Similarity in having any mental disorder (F00-F99) was also considerably higher (r=0.40 vs. r=0.61). Similar estimates calculated from the subsample in combination with median distances between partners’ last place of residence indicate physical proximity and the local pool of potential partners affecting partner similarities.

## Discussion

Using nationwide Finnish registry data on over 1.5 million partnerships, we observed that physical proximity affects partnering in as partners often come from nearby areas. This was more pronounced in more urban municipalities where median distance between partners last place of residence before cohabitation was 12km compared to their rural counterparts where median distance was 56km. Accounting for the local pool of potential partners slightly increased similarities in mental disorders (up 2.4%) but substantially attenuated it for prepartnership educational attainment (11% down), highest obtained degree (22% down) and for 9^th^ grade GPA (24% down), pointing to geographical sorting and spatial clustering of education related variables contributing to observed partner similarities. When further adjusted for both partners’ pre-partnership educational level, the estimates attenuated on average by 3.5% across the different diagnostic categories, suggesting only marginal indirect assortment via education on mental disorders. Higher urbanicity was associated with slightly higher similarity in educational attainment, but slightly lower similarity in any mental disorder, schizophrenia spectrum disorders and behavioral syndromes associated with physiological disturbances and physical factors, consistent with the hypothesis that urban institutions stratify partnering along educational rather then health-related lines. Lastly, within cities, geographic stratification did not affect observed partner similarity. This suggests that geographic barriers may not affect partnering patterns within more densely populated and easily accessible locations. Overall these findings highlight the role the environment plays in partnership formation and suggest different partnering dynamics depending on level of urbanization.

In agreement with previous studies, we observed considerable partner similarities in mental disorders, adding to the growing body of evidence from register based studies ^6,7^, as well as survey based findings ^2,4,5^. Partner similarities in most diagnostic subchapters were in similar range, but we also observed notable variation, e.g. schizophrenia spectrum disorders and intellectual disabilities showing notably higher partner similarities, hinting that assortment may be stronger for some conditions. We observed slightly lower partner correlations in educational attainment than those reported from Norway (Our estimate = 0.39 vs. 0.48 from national registries ^9^ and 0.42 from cohort study ^29^ in Norway), and notably smaller partner correlations for 9^th^ grade GPA (0.20 vs. 0.42 observed in Norway). Educational attainment is challenging to measure especially in young adults who are likely to form partnerships before completing degrees. Thus, we included the highest obtained degree by the end of 2023 to assess if partner similarities would differ between completed degrees and factors underlying the trajectory. Estimates for the highest obtained education by 2023 were slightly larger than for pre-partnership educational attainment (r=0.43 vs. r=0.39), suggesting that partners resemble each other in the latent factors underlying educational trajectories. Alternatively, the difference may reflect age differences between partners and the fact that cohabitation often begins before both individuals have completed their education.

### Accounting for the local pool of potential partners affects observed partner similarity

When municipality-level demographic composition was accounted for, partner similarities in mental disorders were on average 2.4% higher than whole-population estimates, whereas similarities in educational attainment were substantially lower (11–24% depending on measure). This could indicate geographic stratification: mental disorders are somewhat more prevalent in rural areas, whereas more urban areas have higher proportion of highly educated residents ^30^ also observed in our sample of couples (supplementary Table S1). Nationally estimated similarities thus partly reflect regional variation in these traits rather than purely preference-based assortment. Regional variation affected most notably for pre-partnership educational attainment, highest obtained education, and 9^th^ grade GPA, though the observed partner similarities were still substantial after regional variation was accounted for. Earlier research has shown that assortment on educational attainment is stronger than would be expected under direct assortment alone ^11,14^, and previous findings on kin resemblance have found that some of the similarity may stem from residential area ^11^. Our results indicate that geographic clustering of traits, especially for educational achievement, contributes to this pattern.

### Only minimal indirect assortment on mental disorders via education

Indirect assortment on mental disorders would mean that because mental disorders and education are correlated within individuals, assortment on education would also induce correlations between partners in traits that are correlated with education. We observed only minimal indirect assortment on mental disorders contrary to earlier findings (Torvik et al. ^9^. In absolute terms attenuation was similar, as we observed median attenuation for mental disorders of 0.015 (from 0.36 to 0.35) while Torvik et al. observed attenuation for the median values of 0.03. Partner similarities in our study were markedly higher, possibly owing to the sample used, and thus a large proportion of the similarities in mental disorders remain unexplained even after accounting for education. Even if adjusting for both partners educational attainment had attenuated partner correlations in mental disorders, this would not allow to differentiate between preference-based assortment, i.e. shared preferences for higher (or similar) levels of education and social stratification, that is, interactions between people being structured by educational achievement. Whether partner similarities in mental disorders reflect individual preferences for health ^31^, or byproducts of other traits assorted on remains unknown. Based on our findings indirect assortment via education seems minimal.

### Urban and rural areas differ in partnering patterns

Our findings also point towards differences in partner similarity between rural and urban regions. In areas with higher urbanicity, partners were more similar in educational attainment but less similar in some mental disorder diagnoses. These patterns were observed beyond those that would be observed if mating was random within the municipality, and differences remained similar even after adjusting for indirect assortment through education. While a recent study found that partner similarity in mental disorders is a global phenomenon with broadly similar patterns between Taiwan and the Nordic countries Fan et al. ^6^, our findings suggest that meaningful variation may exist within countries.

Our findings provide support to the notion that regional characteristics associated with urbanicity can alter partnering dynamics. Previous literature has identified that in urban settings partners are more similar regarding education ^27,28^ and that partner similarities in religion are affected by local demographic composition ^32^. We extend these previous findings on education to a different context, with larger and more representative data, and with more accurate information on when and where the partnership was formed. Moreover, the pattern was observed when urbanicity was measured on a continuous scale, indicating a gradual urban–rural gradient rather than a dichotomy. The urbanicity classification we used is a holistic measure based on factors such population size, density, and employment related factors, and we were unable to identify if specific features of regions would be particularly important for partnering dynamics.

We also observed a decline in similarity in mental disorders with higher urbanicity. The mechanisms behind this observation remain unknown. It could be that partnering dynamics in rural and urban areas differ. In urban regions partnering may be based on social institutions such as schools and universities ^22–24^, and the role of mental disorders could be smaller in mate selection.

### Physical proximity does not affect partner similarities within cities

Lastly, we observed no differences in partner similarity in mental disorders or educational attainment when limiting the pool of potential partners to within-city subregional units. We also found no differences between more densely populated and less densely populated subregions. Earlier studies have shown that partners often reside close to each other prior to union formation in both the United States ^19^ and the Netherlands (Haandrikman et al., 2008), and we also observed a median distance of 12km in most urban municipalities. Social institutions such as universities, workplaces, and social networks may influence where people live and interact, leading to spatial clustering of traits. However, within cities the costs associated with physical distance, such as travel time, may become distorted, such that physical distance no longer maps linearly onto travel time. Public transportation systems and urban infrastructure can substantially reduce effective travel time within cities, while telecommunications allow individuals to maintain frequent contact regardless of physical distance. Consequently, subregional geographic units within large cities may not meaningfully capture the structure of the local pool of potential partners or the relevant social subpopulations that shape partner selection.

### Integrating regional differences to partnering dynamics

Overall, our findings highlight the role of geography in partnering and in the processes leading to partner similarity. Generally, observed partner similarity in both mental disorders and education was affected by the composition of the local pool of potential partners. Most importantly, our findings strengthen the notion that physical proximity affects partnering in modern societies, but when more densely populated regions are concerned, geographical proximity is less important.

Extending these findings to intergenerational processes, partner similarity is likely intertwined with multiple societal mechanisms. Educational and employment opportunities are among the primary drivers of migration ^33^ and highly educated individuals tend to concentrate in larger cities ^34^, as do genetic predispositions related to education ^35,36^. Such selective migration may increase homogeneity in both origin and destination regions, thereby shaping the local demographic composition and partnering dynamics. The availability of educational and employment opportunities attracts internal migration, reinforcing the spatial concentration of human capital. These dynamics may have intergenerational consequences: children are more likely to grow up in environments with greater educational opportunities, surrounded by peers with similar aptitudes, and within families that possess more resources and knowledge to support educational attainment. In addition, increasing urbanization may lead to stronger social stratification, particularly along educational lines. Although urban populations may be more heterogeneous, individuals in them may be more likely to interact and form partnerships within socially similar groups, as interactions are structured by education, work, and social networks. Over multiple generations, the combination of selective migration and partner similarity may contribute to the accumulation of socioeconomic and health-related advantages within families and within regions, reinforcing existing inequalities, as has been hypothetized earlier ^37^.

### Limitations

Our findings should be interpreted within some limitations. First, we defined partners based on marriage and cohabitation, which allowed us to more accurately pinpoint where and when partnerships began, as couples are more likely to reside together before having children or getting married. However, intergenerational transmission may not be affected to the same degree as only partnerships that result in childbearing contribute to genetic transmission. Consequently, estimates based on cohabiting couples may capture mating patterns that are not fully reflected in the genetic composition of the next generation ^38,39^. Earlier studies from Norway ^9^ and Sweden ^8^ defined couples by shared children and marriage. While defining partnership by shared children is more in line with intergenerational transmission, it may introduce selection bias. People with severe mental disorders are less likely to marry and have children ^40,41^. Furthermore, partnering and childbearing can be partly separate processes and partnering dynamics could be similar regardless of whether the couple has children in the future. Restricting the sample to married couples or parents may disproportionately exclude individuals with psychopathology.

Second, cohabitation was based on official registries (when a couple officially moved together), and we cannot know if the couple met while they were living in different municipalities, or whether partners cohabited before officially moving together. To mitigate this, we performed a sensitivity analysis with only those couples in which both partners had resided within the same municipality for at least two years. Results were broadly similar, and combined with the observed median distance of couples prior to cohabitation, this is more likely to be a problem for rural municipalities where partners lived further apart prior to relationship formation.

Third, using a continuous scale to measure urbanicity allows for more fine-grained analyses compared to dichotomous metrics, and the measure captures multiple dimensions of urbanicity, e.g., population density, population size, and spatial aspects. However, this measure does not capture the specific regional characteristics that affect partnering, and it remains unclear what specific features of the environment, if any, affect partnering dynamics. Fourth, the urbanicity measure reflects Finland specifically. This is also a strength as we were able to include smaller, less densely inhabited municipalities to broaden the view of how environment affects partner similarity. However, the most urban settings in Finland are modest in size by international standards, and our findings may not generalize to bigger metropolitan areas. Fifth, mental disorder diagnoses were acquired from health care registers and although both primary and secondary health care data were available, persons who did not seek help or were diagnosed in primary care before 2011 were classified as not having a mental disorder diagnosis. Validity of mental disorder diagnoses have been reported to be good ^42^, but not all diagnoses have been validated.

Sixth, disentangling contextual effects from temporal effects was not possible with the used spatial scale (municipalities). Our sample included all partnerships formed between 1990 and 2023. In the presence of broader temporal trends (e.g., decline in assortative mating on mental disorders or increasing assortment on education) and increasing migration toward urban centers, the observed urban–rural differences could partly reflect cohort or period effects rather than purely contextual differences. That is, if assortative mating patterns have changed over time and partnership formation has become increasingly concentrated in urban areas, the estimates from urban municipalities may reflect more recent partnerships, whereas estimates from rural municipalities may reflect partnerships that happened earlier in time. An earlier study from Taiwan did observe partner similarities in mental disorders to be rather stable phenomena ^6^, but increasing similarity in educational attainment is well-documented ^24,25,28^. With our design we were unable to answer whether observed urban–rural differences are a result of temporal trends, or whether temporal trends are affected by partnership formation being more clustered to larger cities.

## Conclusions

In conclusion, this study provides evidence for geographic stratification in explaining some of the observed partner similarity in educational attainment but not partner similarity in mental disorders. We also observed differences between regions, with more urban municipalities showing higher similarities in educational attainment, and slightly lower similarities in most mental disorders conditions, compared to their more rural counterparts. This dynamic was present even after adjusting for partners’ educational attainment. Lastly, when using a partner similarities within cities, accounting for subregional units did not affect the observed estimates. This indicates that physical proximity may be relevant only across larger geographic regions, but it becomes unimportant within cities where population density is higher, and where it is easier to meet people who do not necessarily live that close. These findings add to the more complex interactions between an individual and their surrounding environment when modelling partner similarity and emphasize the importance of integrating broader societal context into models of assortative mating.

## Methods

We used nationwide Finnish administrative registers to identify all Finnish residents who started cohabiting between January 1^st^, 1990 and December 31^st^, 2023, and were living within the same municipality when the cohabitation/marriage started. Analyses were limited to different-sex couples, as the coverage of same-sex couples was incomplete. If a couple cohabited/was married more than once, the first available partnership formation date was used. If a person had multiple partnerships, all available partnerships were included. Individual-level register linkages were conducted using personal pseudonymized identity numbers, which are assigned to all Finnish residents.

### Data sources

Data on sex, education, partnerships and place of residence were retrieved from the Full Finnish Population Register (FOLK) of Statistics Finland. For partnerships, the dataset includes the start and end date of cohabitations and marriages. Statistics Finland uses the following definition for cohabitation: a) two individuals of different sexes and over the age of 18 who live in the same residence, are no more than 16 years apart in age, and are not siblings ^43^. Shared children are assumed to mean partnership ^43^.

Information on healthcare contacts was retrieved from the Finnish Care Register for Health Care and from the Register of Primary Health Care Visits. Psychiatric inpatient care has been reliably recorded since 1975, specialist outpatient care since 1998, and primary care data have been available since 2011 ^44^.

In Finland, the International Statistical Classification of Diseases and Related Health Problems, Tenth Revision (ICD-10), has been in use since 1996. Earlier diagnostic coding was based on the Finnish adaptation of ICD-9 between 1987 and 1995, and ICD-8 between 1969 and 1986. In certain primary care settings, diagnoses are recorded using the International Classification of Primary Care, Second Edition (ICPC-2), rather than ICD-10. These diagnoses were harmonized by converting them into the corresponding ICD-10 subchapter categories ^45^, and the register data were further processed to enhance data accuracy ^44^.

GPAs were obtained from the National Joint Application Register containing information on school grades of students in the final year of comprehensive school since 1996. Between years 1996–2007 information on grades was available only for those who applied to secondary education. From 2008 onwards information was available for all students in the final year of compulsory school. The GPA was calculated based on all subjects including compulsory (e.g., mother tongue and literature (Finnish or Swedish), mathematics, history and social studies, music, and visual arts) and optional (e.g., the second foreign language) subjects.

The ethics committee of the Finnish Institute for Health and Welfare approved the study plan (THL/184/6.02.01/2023§933). Data were linked with the permission of Statistics Finland (TK-53-1696-16) and the Finnish Institute for Health and Welfare. Informed consent is not required for register-based studies in Finland. The Strengthening the Reporting of Observational Studies in Epidemiology (STROBE) reporting guideline was followed.

### Measures

Mental disorder diagnoses (ICD-10 F10 to F99 and equivalent ICD-8, ICD-9, and ICPC-2 codes), included inpatient hospital episodes, secondary outpatient visits including emergency department visits, and primary healthcare visits in Finland. We used the following ICD-10 subchapter F categories: any mental disorder (F00-F99), substance use disorders (F10-F19), schizophrenia spectrum disorders (F20-F29), mood disorders (F30-F39), anxiety disorders (F40-F48), behavioral syndromes associated with physiological disturbances (F50-F59), personality disorders (F60-F69), intellectual disabilities (F70-F79), developmental disorders (F80-F89), and childhood onset disorders (F90-F98). A person was considered diagnosed if they received their diagnosis prior to the start date of the cohabitation/marriage and undiagnosed if they had no record of a diagnosis or the first diagnosis occurred only after the start of cohabitation/marriage.

We used three measures of educational attainment, 1) highest obtained degree when partnership began (pre-partnership educational attainment), 2) highest obtained degree by the end of 2023, and 3) grade point average at the end of 9^th^ grade. Educational attainment was measured on a 7-point scale ranging from 1 = only compulsory education to 7 = Doctoral degree. The GPA was standardized by birth year (mean = 0, sd = 1) and available for 215 192 men and 340 894 women.

### Municipalities and urban–rural classification

Finland has 309 municipalities (2024 classification), which constitute the lowest level of administrative governance and are responsible for core public services. Municipalities in the Åland archipelago were combined, due to their small size and isolated location, giving a total of 293 municipalities. In the main analysis, a couple’s municipality was determined as the municipality where first cohabitation or marriage occurred. To measure urbanicity, we used the seven-level urban–rural classification for the year 2010 issued by the Finnish Environment Institute based on a nationwide grid of 250 × 250m cells ^46^. The seven levels are 7 = inner urban areas, 6 =outer urban areas, 5 = Peri-urban areas, 4 = rural local centers, 3 = rural areas close to city, 2 = core rural areas, and 1 = sparsely populated rural areas. The classification is based on population density, built environment, commuting links, and land-use characteristics. Each municipality’s urbanicity was assigned based on the urban–rural level in which most of its inhabitants resided.

In within-city analyses we analysed three cities, Helsinki, Turku and Tampere separately using postal code areas. These cities were divided into subregions shown in Figure 4, supplementary Figure S2 and supplementary Figure S3, to investigate if there are within-city differences in partnering.

### Statistical analysis

We calculated the prevalence of each examined ICD-10 category and means and standard deviations of education-related variables at individual level before the start of marriage or cohabitation. Next, we estimated the degree of partner similarity in mental disorders with tetrachoric correlations within diagnostic categories. For education related variables, we used Pearson correlations to estimate partner similarity in the pre-partnership educational attainment, highest obtained degree and 9^th^ grade GPA. We then calculated partner similarity within each diagnostic category, and for each educational variable stratified by municipality. The Idea here was that if people are more likely to meet their partner within close physical proximity, and regions differ in their demographic composition, the potential pool of partners differs between municipalities and partner similarities may arise without individual preferences. If the whole population is used to calculate partner similarity, random mating would mean that any two people of different sexes could partner regardless of the location. When partner similarity is calculated within a municipality, the assumption is that people would find partners within the same municipality. While this does not always hold, given the close physical proximity between partners before cohabitation (Fig. 1), and the 31.5km median distance between neighbouring municipalities in Finland, using municipalities offers a practically meaningful way to model the pool of potential partners.

As some of the smaller municipalities did not have enough cases to calculate tetrachoric correlations, we tried estimating tetrachoric correlations for each municipality and used those where estimation was successful and reasonable estimate was observed. The number of municipalities used in each analysis is reported in supplementary Table S3. We then aggregated municipality-specific partner correlations using random-effects meta-analysis by first transforming the correlations with Fisher z-transformation and after meta-analysis backtransforming the estimates to original scale. For Polychoric correlations standard errors were transformed using delta method. Lastly, we adjusted municipality-specific estimates of partner similarities on mental disorders for both partners’ pre-partnership educational attainment similar to Torvik et. al ^9^, and aggregated these using random-effects meta-analysis.

### Meta-regression

Urban–rural differences were studied with meta-regressions, with urbanicity as a fixed-effect moderator, treated as continuous variable, for municipalities. Correlations were transformed with Fishers z-transformation, and standards errors for polychoric correlations were transformed using delta method. These analyses were done with municipality-specific estimates for mental disorders and education, and with municipality-specific estimates adjusted for education for mental disorders. All estimates and their confidence intervals were obtained via restricted maximum likelihood.

### Within-city analyses

In a similar fashion, we analysed partner similarities in three major cities comparing estimates calculated from all partnerships within the municipality, to estimates aggregated from subdistrict-specific estimates. We also tested population density as a potential moderator using meta-regressions with population density as a fixed-effect moderator.

### Controlling for multiple comparisons

In each step we had 13 comparisons (9 mental disorders subchapters, 1 indicator of any mental disorder, 3 education variables (current EA, highest obtained EA, 9^th^ grade GPA). We did analyses in three sets, first comparing whole-sample estimates to aggregated municipality-specific estimates, second, effects of urbanicity, and third within city analyses. Hence, we corrected the p-values with a set significance level at .05, which was then adjusted with a Bonferroni correction.

### Sensitivity analyses

We also conducted a sensitivity analysis to evaluate the robustness of the findings. As the relationship formation may not happen when partners are living in same municipality, or it may happen so that both partners move to a new location when cohabitation starts, we analysed a subsample where both partners had lived within the municipality where cohabitation/marriage started for at least two years.

Data management was performed using R version 4.6.0 ^47^ and Stata version 17.0. Analyses, figures, and tables were generated in R version 4.6.0 ^47^ using the packages lavaan ^48^, polycor ^49^, metafor ^50^, and tidyverse ^51^.

## Data availability

Data used in this study are the property of Statistics Finland and the Finnish Institute for Health and Welfare. The data are available from the respective authorities, but certain restrictions apply. For more information on accessing the data, please visit www.findata.fi.

## Acknowledgements

The authors thank Vilja Helminen for valuable feedback during the early stages of this work and for her assistance with the visual elements presented in this paper. The present study was funded by the European Union (ERC, MENTALNET, 101040247) and the Research Council of Finland (354237 to CH). ES was funded by the Research Council of Finland (364371,364384). KS was funded by The Finnish Medical Foundation (9198). Views and opinions expressed are however those of the authors only and do not necessarily reflect those of the European Union or the European Research Council. Neither the European Union nor the granting authority can be held responsible for them.

## Contributions

E.S conceived the idea and designed the study, with support from C.H.

E.S, M.G, and K.K contributed to data curation.

E.S carried out the analyses and visualised the results with support from C.H.

K.G, K.K, , M.G, M.E, and M.J contributed to interpretation of the results.

E.S wrote the manuscript with input from all authors. All authors provided critical feedback, discussed the results, helped shape the manuscript, and approved of the final manuscript.

